# Development and scaling of a sequencing pipeline for genomic surveillance of SARS-CoV-2 in New York City

**DOI:** 10.1101/2022.05.25.22273991

**Authors:** Michael J. Hammerling, Shinyoung Clair Kang, William Ward, Isabel F. Escapa, Pradeep Bugga, Cybill Del Castillo, Melissa Hopkins, Steven Chase, Sol Rey, Dylan Law, Alexander Carpio, Katharine Nelson, Simran Chhabria, Simran Gupta, Tiara Rivera, Jon M. Laurent, Haiping Hao, Henry H. Lee

## Abstract

In the ongoing COVID-19 pandemic, detecting the appearance and spread of variants of concern (VOC) is a critical capability in the fight to quell the virus and return to normalcy. Genomic surveillance of the emergence, propagation, and geographical spread of VOCs is thus an important tool for public health officials and government leaders to make policy decisions and advise the public. As part of our role as a major SARS-CoV-2 diagnostic testing facility in New York City, the Pandemic Response Lab (PRL) has been performing genomic surveillance on the large number of positive samples processed by the facility on a daily basis from throughout the New York metropolitan area. Here we describe the development and optimization of a high-throughput SARS-CoV-2 genome sequencing facility at PRL serving New York City.

## INTRODUCTION

The COVID-19 pandemic has placed an unprecedented burden on healthcare institutions across the world and revealed a lack of infrastructure capable of scaling quickly to counter pandemic-scale public health crises. In the United States, New York City was the most severely impacted metropolitan area early in the pandemic, with the 7-day average number of new confirmed cases reaching 5,132 per day in late March 2020^1^. Coupled with an inadequate diagnostic testing infrastructure, this early surge led to a scarcity of testing capacity, necessitating a total lockdown as the only available measure to fight the pandemic. As a first step to allowing the city to reopen, a home-grown diagnostic facility was needed to mitigate the economic and public health crisis and provide testing capacity for New York City.

The Pandemic Response Lab (PRL) was established in September 2020 as a Manhattan facility completely dedicated to meeting the SARS-CoV-2 testing needs of New York City and its five boroughs. This facility uses a proprietary and scalable automated real-time RT-PCR based testing pipeline developed by NYU Langone Health scientists, which is currently capable of processing over 45,000 tests per day with turnaround times under 24 hours. This increased capacity has been instrumental in enabling the public health apparatus of New York City to meet the testing needs brought on by the multiple waves of SARS-CoV-2 cases that have impacted the city over the course of this pandemic.

Despite the success of this increased testing infrastructure, new variants of SARS-CoV-2 have continually emerged throughout the pandemic, supplanting earlier variants and seeding new surges in cases. Concerns over the evolution of viral variants prompted calls to perform whole-genome sequencing on SARS-CoV-2 genomes to catalog and track their lineages. To date, the most significant variants of concern (VOCs) are Alpha (B.1.1.7), Beta (B.1.351), Iota (B.1.526), Gamma (P.1), Delta (B.1.617.2) and most recently, Omicron (B.1.1.529, BA.1, BA.2, or BA.3), primarily due to their increased infectivity^2–5^, higher viral load^6^, or potential for immune evasion^7,8^. To understand the changing dynamics of VOC proliferation in NYC, a citywide genomic biosurveillance system was proposed to track the emergence and spread of VOCs for the purpose of informing policy decisions and implementing public health countermeasures.

Beginning in January 2021, the PRL R& D facility in Long Island City (LIC) implemented a high-throughput processing pipeline to sequence positive cases of SARS-CoV-2 detected by our testing facility in Manhattan (now also located in LIC). Because the end-to-end process of swab receipt, diagnosis, and genome sequencing can be vertically integrated at our facilities, this pipeline is able to routinely offer turnaround times for sequenced genomes of confirmed positive cases within 3 days of receipt at the sequencing lab. Our final pipeline entails “hitpicking”, or automated aggregation of SARS-CoV-2 positive remnant samples into new plates at our clinical diagnostic facility, followed by transport of hitpicked samples from the diagnostic lab to our R& D facility for genome RT-PCR, library preparation, and sequencing (**Figure 1**). We also established a parallel workflow (STATseq) to directly accept specimens diagnosed as positive for SARS-CoV-2 virus by various external testing platforms, most notably from the Emergency Department of Health and Hospital Centers (HHC), for genome sequencing. RNA from these samples is extracted in our R&D facility and fed directly into our sequencing pipeline, enabling sequencing as fast as within a day of sample receipt.

**Figure 1.**
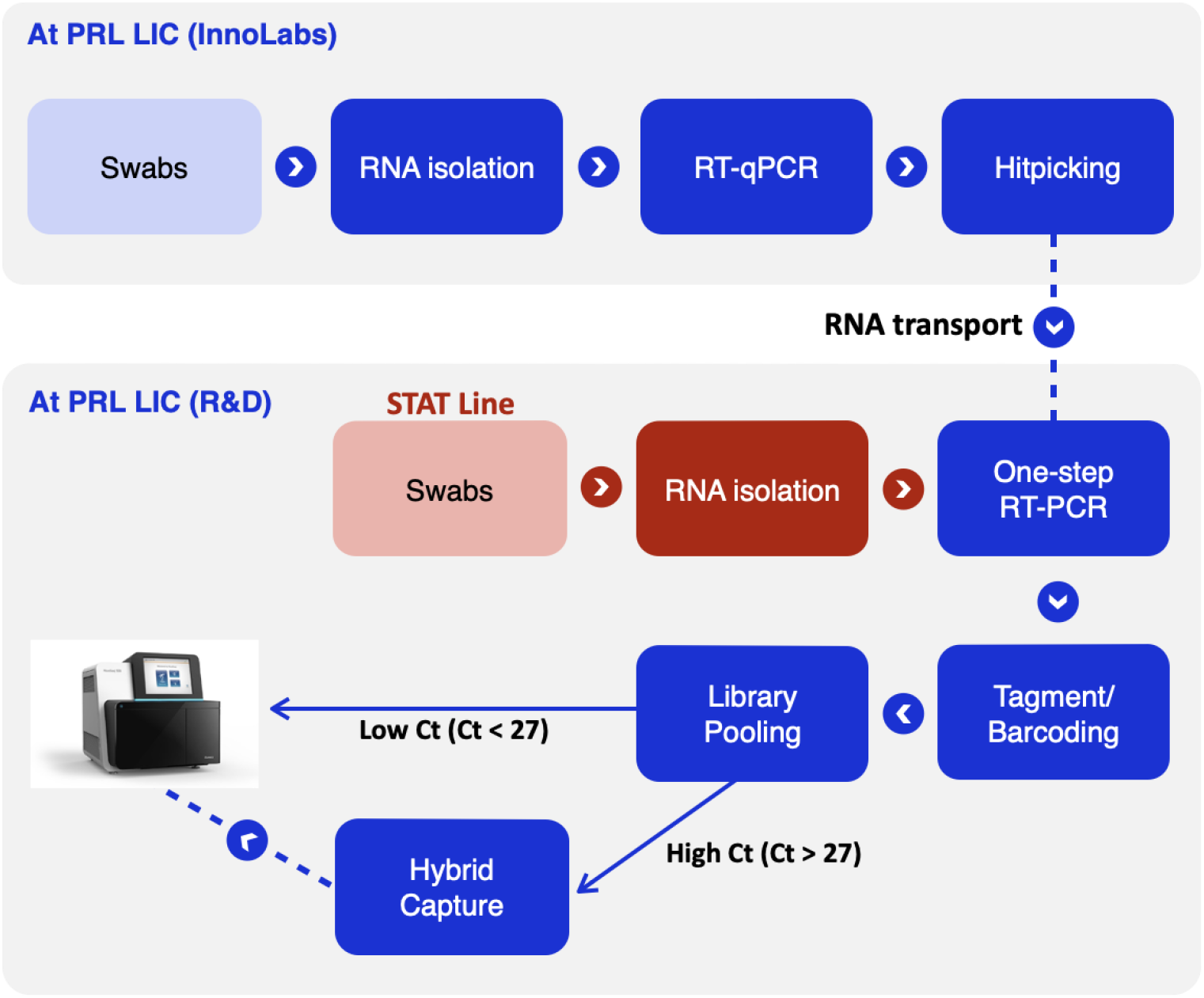
PRL integrated pipeline for COVID-19 testing and sequencing. The integrated pipeline begins with RNA extraction and RT-qPCR-based testing of incoming patient samples at the InnoLabs facility. After categorizing samples as either PCR-positive or negative, residual RNA from samples which are positive for SARS-CoV-2 RNA are automatically identified and reformatted, or “hitpicked”, into 384-well plates for processing in our sequencing pipeline. The positive samples are then converted to DNA using multiplexed RT-PCR, and tagmentation-based library prep and barcoding is performed on the resulting cDNA. Samples are then pooled separately into low Ct and high Ct bins, and the samples in the high Ct bin go through an extra step of hybrid capture before being sequenced on an appropriate Illumina platform. A parallel workflow to directly accept specimens from external facilities (the STATseq line) was also established. Specimens diagnosed as positive by the external platform are extracted in the R& D facility and fed directly into the sequencing pipeline. See Materials and Methods for more detail on all steps of the pipeline.

To date, our facility has generated >75,000 SARS-CoV-2 genomes for surveillance and research purposes, and deposited >54,000 genomes to GISAID^9^. With this pipeline, we have been able to identify the earliest introduction of Beta (B.1.351), Gamma (P.1), Delta (B.1.617.2), and Omicron (B.1.1.529) at ports of entry (JFK airport) or via community spread. Here we document the process improvements, optimizations, and automation of this scalable, high-throughput, and low-cost sequencing pipeline, and its contribution to metropolitan-wide surveillance of the spread of VOCs in New York City. Where possible, we discuss trade-offs to maximize scalability while preserving the data quality critical for epidemiological study and resulting public health measures.

## RESULTS

To maximize the biosurveillance capabilities of our COVID sequencing pipeline, we describe below process improvements tested and implemented in our pipeline which improved reconstruction of viral genomes across the range of patient viral loads (i.e. observed cycle threshold (Ct)), sample turnaround time, and pipeline scalability.

### 1. RNA quality control

Preservation of RNA sample quality before library preparation for sequencing is the crucial first step of the sequencing pipeline. We observed that RNA degradation caused by even short times at ambient temperature or freeze-thaw cycles, as may occur during transport, storage, or hitpicking, dramatically reduces the reconstruction success rate for SARS-CoV-2 positive samples. To investigate the degree of sample degradation during transport, RT-qPCR using both the CDC N1 and N2 primer-probe sets (CDC EUA200001) was performed after transport and hitpicking at our sequencing facility and compared to those reported at the PRL testing facility. For samples with delays in transport, degradation is detected as an upward shift in Ct value compared to those reported at the PRL testing facility (**Supplementary Figure 1A**). This is true especially for samples with low RNA concentration or degraded RNA (high Ct). To avoid these issues, we implemented a strictly regimented process for temperature control during shipping, storage, and hitpicking to preserve RNA quality between the diagnostic lab and sequencing lab facilities (**Materials and Methods**). Following the implementation of improved logistical standards, Ct values for both primer-probe sets were observed to align closely with the original values obtained at the Manhattan testing facility, indicating that these quality control measures are successful in preserving RNA quality for downstream sequencing (**Supplementary Figure 1B**). Regular confirmation of Ct values for a subset of samples from each plate entering the sequencing pipeline was thus implemented as standard operating procedure to detect deviation from expected Ct values that may be caused by delays stemming from a rapid surge in positive cases, such that changes may be quickly observed and corrective measures implemented.

### 2. RT-PCR

#### 2.1 Selection of an effective RT-PCR strategy and enzyme mix

For amplification and library preparation of the SARS-CoV-2 genome, we opted to combine the Midnight primer set and protocol^10,11^, which amplifies the genome in larger 1200 bp fragments across two multiplexed PCR reactions, with tagmentation for library fragmentation and indexing. This strategy reduces the amount of the genome to which primers anneal compared to other primer sets, lowering the likelihood of fragment dropouts due to accumulation of mutations in those primer binding regions. We believe this combination of methodologies is unique among large SARS-CoV-2 sequencing facilities.

The Midnight protocol calls for using a two-step RT-PCR protocol utilizing the LunaScript®RT SuperMix Kit for RT and Q5® Hot Start HF 2x Master Mix for PCR amplification of the cDNA. While the protocol and components were initially implemented as described, we later wanted to explore the substitution of one-step RT-PCR kits to reduce labor, cost, and pipeline runtime, and to improve performance. A variety of 1-step kits were tested on extracted SARS-CoV-2 RNA samples, including the NEB OneTaq One-Step (#E5315S), Invitrogen SuperScript™ IV One-Step (#12594025), Promega GoTaq® 1-step (#A6121), and Takara One Step PrimeScript III (#RR600A) kits. Of the kits tested, the Takara kit with two-step cycling was found to reliably produce the most robust amplicon bands across the broadest range of Cts compared to other one-step kits (**Supplementary Figure 2**).

Considering this, the Takara kit was chosen to move forward with a side-by-side comparison against the two-step Lunascript-Q5 protocol. The Takara one-step kit was often successful in generating the desired viral amplicons where Lunascript-Q5 failed (**Figure 2A**). In a test of 384 RNA SARS-CoV-2 positive RNA samples, the number of genomes reconstructed by the Takara one-step kit was higher than the two-step protocol, allowing reconstruction of 263 genomes compared to 226 genomes out of 384 RNA samples attempted across a wide range of Ct values. Takara also performed better across a variety of other metrics, including yielding fewer uncalled bases (**Figure 2B,** Supplementary Figure 3A-D), greater average genome coverage (**Figure 2C, Supplementary Figure 4A-D**), and more complete genomes (**Figure 2D, Supplementary Figure 5A-D**). This was true even and especially in cases where the genome construction did not meet our criteria for a passable genome (**Materials and Methods**), but still yielded usable genome information (**Supplementary Figure 3D, 4D, and 5D**).

**Figure 2.**
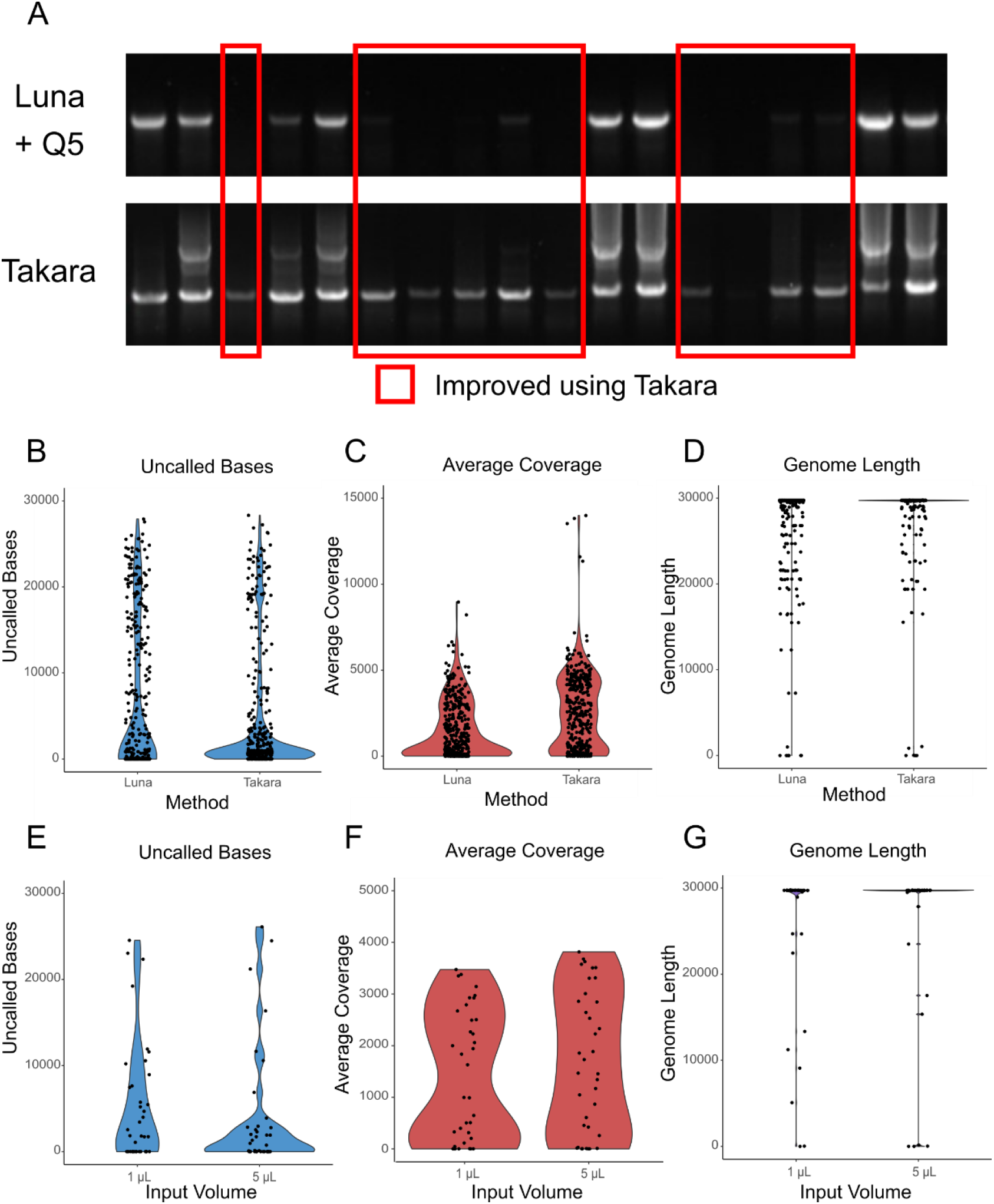
Optimization of RT-PCR kit and parameters for the generation of SARS-CoV-2 cDNA for genome sequencing. **(A)** RT-PCR of 18 SARS-CoV-2 RNA samples was performed using the two-step Lunascript + Q5 protocol or the Takara 1-step kit. For 9 of 18 samples, the 1-step kit performed better at generating a visible band than the 2-step protocol. **(B)** 384 SARS-CoV-2 positive RNA samples were sequenced using both the 2-step and Takara 1-step protocols. In addition to generating more reconstructed genomes, the Takara 1-step protocol generated fewer uncalled bases **(B)**, better average genome coverage **(C)** and longer consensus genomes **(D)** than the 2-step protocol. RT-PCR was also performed on 37 samples using either 1μL or 5μL of sample as input in the reaction. The 5μL reactions were found to have (**E**) fewer uncalled bases, (**F**) greater average coverage, and (**G**) longer average genome length than achieved in the 1μL cases.

While the Takara one-step kit is expected to have a slightly higher error-rate on the nucleotide scale compared to the two-step protocol due to the lack of a high fidelity proofreading DNA polymerase, we opted to use this kit moving forward due to several advantages. For whole-genome reconstruction, we reasoned that the effect of random mutations that occur in amplicons could be mitigated by allocating sufficient sequencing read depth and setting a high threshold for calling mutations when generating the consensus sequence. Given the savings in cost, labor, and time, the greater number of genomes constructed, and the substantially higher average quality and length of the genomes produced with the Takara one-step kit **(Figure 2B-D**), we reasoned that we could improve both our sample throughput and quality of results by converting the sequencing pipeline to utilize this kit moving forward.

#### 2.2 Assessment of over-sequencing of high Ct samples as a strategy to improve genome reconstruction

Samples with low RNA concentration (Ct ≥ 30) often produce low-quality or incomplete genomes without targeted intervention to improve the results of these samples^12–14^.We observed that such high-Ct samples generally produce fewer on-target sequencing reads than low-Ct samples. Consequently, we wanted to test whether greatly increasing sequencing depth of these samples would allow high Ct samples to be reconstructed. Seven samples with N1 Ct values between 29 and 40 were prepped and sequenced along with a single low Ct sample (Sample 2, N1 Ct = 16.45) to control for any possible errors in library preparation of the high-Ct samples. Sequencing libraries for these samples were made equimolar before being loaded on NextSeq using Illumina NextSeq 500/550 Mid-Output v2.5 Kit (150 cycles). Even with these high Ct samples receiving as many as 1.3 million reads, the samples still failed to reconstruct with the number of uncalled bases reaching 17,000 or higher (out of 29,870) and low overall coverage (**Supplementary Table 1**). This result demonstrates that increasing sequencing depth alone cannot compensate for the relatively poor quality of high Ct samples to rescue genome reconstruction. Instead, the failure of an overabundance of sequencing coverage to save these samples implicates factors influencing the conversion of viral RNA to a complete complement of cDNA, such as quality and quantity of source RNA, RT-PCR parameters, and the quality of the tagmented libraries as more crucial in determining whether the entire genome is represented in the sequencing library. For this reason, much of the optimization efforts described in the following sections were focused around improving these parameters.

#### 2.3 RT-PCR cycling parameters and input volume

To further improve pipeline performance while reducing protocol runtime, we sought to optimize the PCR cycling parameters. First, a temperature gradient was performed during the elongation cycle of the 2-step RT-PCR cycling protocol utilizing the Takara PrimeScript kit to determine the optimal extension temperature. The extension temperature of 61°C was found to be optimal for producing an RT-PCR band from the most samples while minimizing off-target product formation (**Supplementary Figure 6**). We next sought to minimize the extension time without compromising the ability to produce products from marginal, high Ct samples. We found that extension times less than 3 minutes resulted in loss of genomic PCR product from some samples (**Supplementary Figure 7**), while extension times longer than 3 minutes resulted in the formation of large molecular weight off-products. These findings were incorporated into the final two-step cycling protocol for RT-PCR of the SARS-CoV-2 genome (**Materials and Methods**).

Next, the volume of sample RNA added to the RT-PCR reaction was optimized. While increasing total RNA in the reaction should intuitively improve RT-PCR performance of low-concentration samples, residual contaminants from the RNA extraction procedure can also inhibit RT-PCR. Therefore, it was necessary to determine the RNA volume that would maximize the RNA input without compromising RT-PCR performance. When tested on STAT samples that were directly received and extracted at the LIC site using Kingfisher Flex (Thermo Scientific), there was a subtle change in the number of uncalled bases, average coverage, and genome length for different input volumes, and best results were observed with higher input volume of 5 uL (**Figure 2E-G, Supplementary Table 2**). However, changes to extraction procedure such as differing automation methods, elution volumes, or sample handling/timing can change the outcome of differing sample input volumes by virtue of altering the purity and concentration of the eluted sample. Consequently, this parameter should be optimized for any new genome sequencing operation which may use alternative extraction methods or instruments.

#### 2.4 Optimization of A1200 primers

Multiplexed PCR reactions may suffer from uneven amplicon representation due to variable strength of annealing of primers to template, formation of specific primer dimers, inherent instability of genomic regions, and other factors^15^. In our pipeline, SARS-CoV-2 positive samples which were sequenced using equimolar concentrations of the Midnight primers in the genome amplification RT-PCR reaction resulted in uneven amplicon representation measured as the fraction of sequencing reads aligned to that amplicon region (**Figure 3A and 3B**). In particular, amplicons 5, 15, 21, and 23 from primer set P1 and amplicons 6 and 20 from primer set P2 have consistently low representation in final sequenced libraries. Dropout of these amplicons is often responsible for failure to reconstruct a complete genome for positive samples. Rebalancing of primer concentrations in the multiplexed reaction is a proven strategy to remedy uneven amplicon distribution in multiplexed PCRs^16^, and has been used in sequencing of SARS-CoV-2 genomes to mitigate these issues and make amplicon representation in sequenced pools more even^17^. Furthermore, mutations in the annealing region of the original Midnight primer set can disrupt primer binding and lead to amplicon dropout. In particular, the Omicron variant has mutations in the binding site for primer pairs 10, 24, and 28 which can significantly impact the ability to sequence these regions^18^.

**Figure 3.**
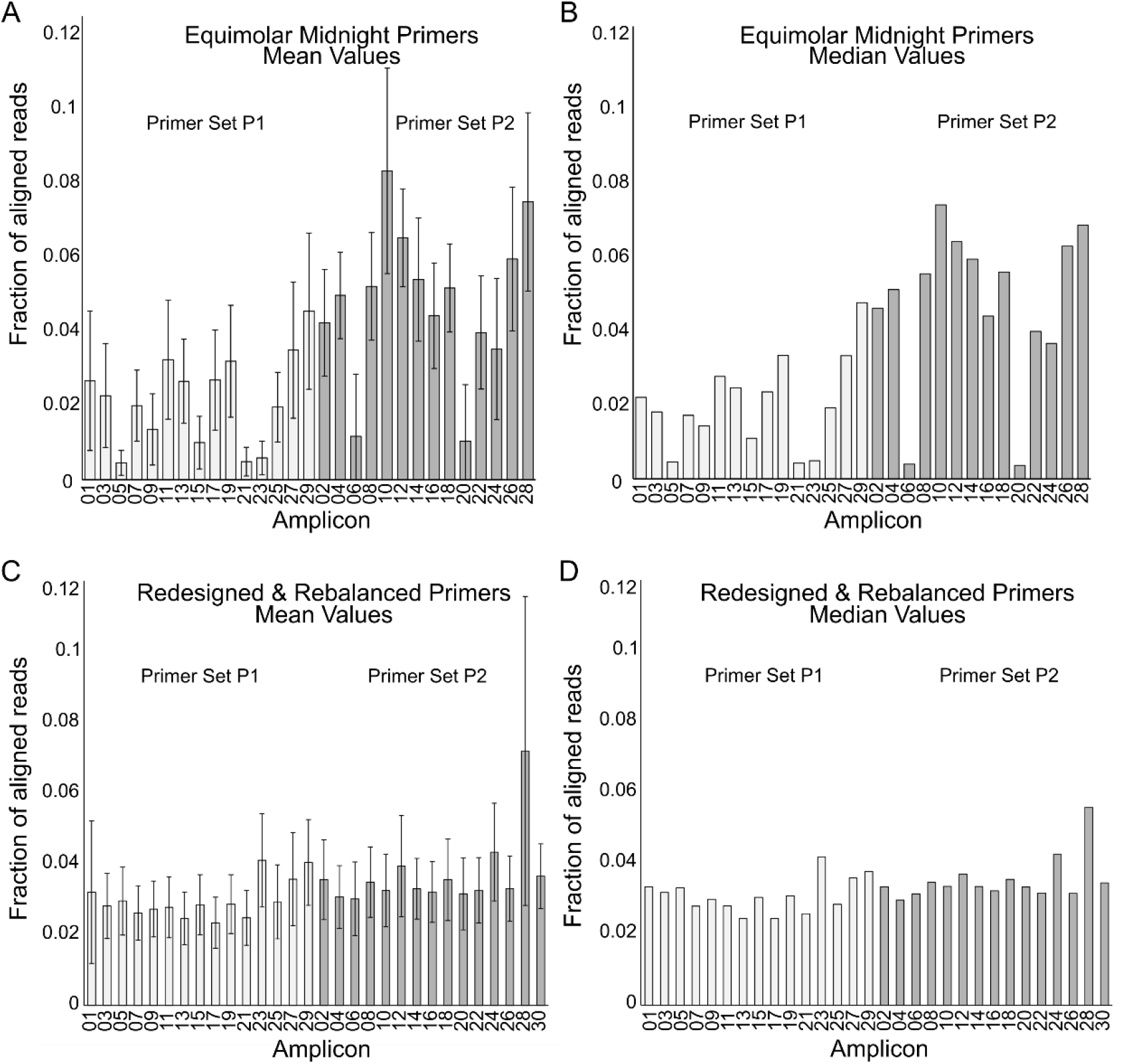
Redesign and rebalancing of A1200 primers for more even representation of amplicons. (**A)** Mean and (**B**) median per sample fraction of aligned reads for each amplicon of the Midnight primer set were measured for 29 samples which were successfully reconstructed using equimolar primer concentrations in May 2021. Amplicons suffer as much as a 11.5-fold difference in representation between the mean values for the highest (A08) and lowest (A05) represented amplicons. Following our most recent primer redesign and rebalancing to adapt to the Omicron variant, (**C**) mean and (**D**) median values for per sample fraction of aligned reads for each amplicon were assessed for 940 successfully reconstructed samples sequenced on April 8, 2022, resulting in a maximum 3.1-fold difference between the mean values for the highest (A28) and lowest (A17) represented amplicons. Error bars represent one standard deviation.

Both redesign and rebalancing of the Midnight primer set was required to mitigate these emerging challenges to genomic surveillance of the Omicron variant. Primer pools P1 and P2 were redesigned to avoid mutations present in the dominant Omicron strain while retaining the overall 1200 bp size target for amplicons, creating the new PRL A1200 primer set, which generates 30 amplicons compared to the 29 in the Midnight primer set (**Supplementary Data Table 1**). To inform rebalancing of primer concentrations in the multiplexed PCR reaction, the representation of each amplicon in successfully reconstructed genomes using equimolar ratios of the Midnight primer set were input into a previously described equation to generate a modified weight for each primer pair in pools P1 and P2^16,17^. These representation values were used to determine the weight of each 100 uM primer to input into the rebalanced primer sets (**Supplementary Data Table 1**). The rebalanced primer pools achieve more even coverage across problematic amplicons (A05, A21, A23, A06, and A20), and rescue amplicons impacted by Omicron mutations (A10, A24, and A28) (**Fig. 3C and 3D**).

### 3. Tagmentation and Barcoding

Since the Midnight protocol produces SARS-CoV-2 cDNA in 1200 bp fragments^11^, but short-read Illumina sequencing is the most cost-effective way to assemble large numbers of SARS-CoV-2 genomes, we needed an rapid and reproducible method for fragmenting our cDNA libraries and attaching indices and barcodes. Tagmentation is a method utilizing a hyperactive mutant of transposase Tn5 which enables simultaneous fragmentation and ligation of indices to SARS-CoV-2 cDNA products in a single, brief isothermal reaction^19,20^. This method dramatically reduces library preparation time compared to ligation-based methods. Following sample tagmentation, identifying barcodes can be added to each sample so they may be pooled and sequenced on a single Illumina sequencing run (**Materials and Methods**). However, as tagmentation mix is one of the most expensive components of the lab pipeline, optimizations were desired to reduce reagent consumption and cost.

#### 3.1 Using polyethylene glycol (PEG) to improve tagmentation efficiency and recovery of high Ct samples

PEG has been shown previously to be beneficial in tagmentation reactions for NGS library construction to promote efficient tagmentation with reduced reagent concentration^19,21–23^. To reduce the cost of library preparation, we wanted to test the ability of PEG to promote tagmentation of A1200 amplicons in our miniaturized reaction using reduced transposome concentrations. To test the efficacy of this strategy, tagmentation was carried out under standard reagent concentration without PEG, and with 8% PEG 8000 and 0.1X the prescribed concentration of transposase enzyme (**Materials and Methods**). We observed a similar or slightly better full genome reconstruction rate for higher Ct samples in comparison to tagmentation without PEG despite the 10-fold decrease in quantity of transposome used (**Figure 4A**). Importantly, the majority of improvement from using PEG was realized on higher Ct samples (**Figure 4B**). These genome reconstructions also did not have a significantly different number of uncalled bases despite the 10-fold decrease in quantity of transposome used (two-sided t-test, p = 0.9692) (**Figure 4C**). Since transposase enzyme is one of the most expensive components of our wet lab sequencing pipeline, this technique provides a way to reduce the cost of tagmentation significantly without sacrificing library quality. This is also an effective strategy to preserve transposase enzyme in instances where supply chain issues make procuring new enzyme in a timely manner challenging, or when increases in sample volume (such as during the Omicron surge) have rapidly depleted our stocks.

**Figure 4.**
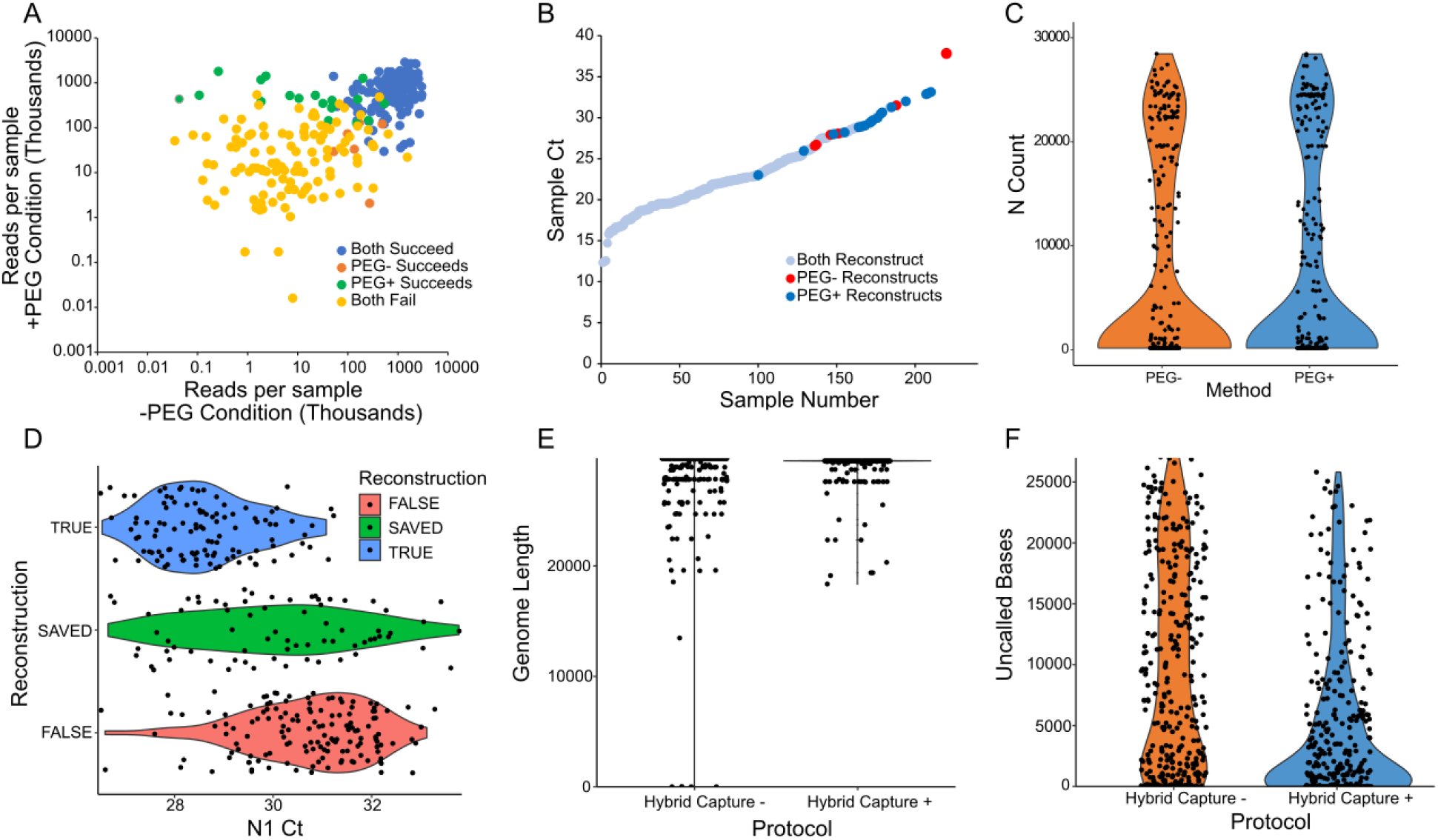
Increasing effective concentration of SARS-CoV-2 cDNA during tagmentation and in the final sequenced libraries improves sequencing outcomes. **(A)** RT-PCR was performed on a set of 288 SARS-CoV-2 positive samples which were then tagmented under standard conditions or with 8% PEG 8000 and 10-fold reduced transposase concentration. Samples tagmented with PEG in general achieved greater reads per sample, and 16 genomes reconstructed only in the PEG+ condition (green), compared to only 6 in the PEG-case (orange). **(B)** Reconstructed samples from this experiment were plotted by Ct and colored by whether genome reconstruction succeeded in both treatments (light blue), only in the PEG-condition(red), or only in the PEG+ condition (dark blue). Samples which reconstruct only in the PEG+ case are highly biased toward high-Ct samples, suggesting that these samples may preferentially reconstruct due to molecular crowding induced by PEG. **(C)** Samples from the PEG- and PEG+ conditions are displayed in violin plots of the number of uncalled bases with individual points represented. There is no statistically significant difference in the overall number of uncalled bases per sample. **(D)** 384 samples above Ct 26.5 were sequenced either with or without hybrid capture of barcoded libraries. Samples which were reconstructed in both conditions are labeled TRUE, samples which were reconstructed in neither are labeled FALSE, and samples reconstructed only when hybrid capture was applied are labeled SAVED. Saved samples are biased toward higher Ct. **(E)** Reconstructed genome length is plotted for both samples to which hybrid capture was and was not applied. Genome lengths are longer when samples are treated with hybrid capture. **(F)** Uncalled bases are plotted for both samples to which hybrid capture was and was not applied. Samples have fewer uncalled bases on average when hybrid capture is applied.

#### 3.2. SARS-CoV-2 DNA concentration normalization before tagmentation

Due to the high number of samples that are processed together in batch and the broad range of RNA input concentrations across samples entering the RT-PCR reaction, it is common to see high sample-to-sample variance in the cDNA concentrations going into tagmentation and barcoding, and thus the number of reads for each sample during sequencing. One strategy to achieve more even coverage of final barcoded libraries is to normalize concentration after RT-PCR, but before tagmentation and barcoding. To test our ability to achieve more uniform read depth for sequencing runs containing many samples, we tested the impact of manual normalization of A1200 amplicons after A1200 RT-PCR and barcoding PCR on sequencing outcome.

For amplicon normalization, the DNA concentrations of A1200 amplicons RT-PCRed from 32 RNA samples in the N1 Ct range of 20 to 32 were measured using a Qubit fluorescence assay (Thermo Fisher). Following amplicon quantitation, the samples were individually diluted to 0.5 ng/uL. The rest of the tagmentation and barcoding steps were carried out as normal. After barcoding PCR, DNA quantification was again performed to obtain the concentrations of barcoded libraries. Based on the measured concentrations, the libraries were made equimolar before library pooling. This normalization method helped achieve a marginally more uniform percentage reads per sample across a large Ct range (**Supplementary Figure 8**). However, due to the limited effectiveness and high labor investment of this intervention, manual normalization of individual cDNA samples before tagmentation was not implemented. Instead, the concentration of a subset of RT-PCR reaction of representative Ct is measured using Qubit reagent, and one of several pre-programmed bulk dilution protocols is implemented to bring concentration of all samples into the <0.5 ng/μL range necessary for tagmentation to proceed efficiently (**Materials and Methods**).

### 4. Hybridization capture of constructed DNA libraries improves sequencing of high Ct samples

Hybridization (hybrid) capture is a method for enriching nucleic acid sequences of interest from samples with high complexity or low concentration using antisense oligonucleotides to achieve more on-target sequencing reads^24^. This approach is useful for downstream applications, such as pathogen detection and identification, genomic characterization, and identifying virulence determinants^25–27^.We hypothesized that hybrid capture would be especially useful for improving the reconstruction rate of high-Ct samples, which did not perform as well in the A1200 RT-PCR reactions or downstream steps in the sequencing pipeline.

A hybrid capture technique was applied to improve genome reconstruction of more challenging samples with low viral loads using TWIST SARS-CoV-2 Research Panel (#103567, Twist Biosciences). Barcoded libraries from samples with Ct value above 26.5 from each sample plate were pooled together separately from the low Ct samples and then processed into a single hybrid capture reaction with a total of 0.7-1.5 ug DNA input at a time to enrich for SARS-CoV-2 sequences. Application of hybrid capture to high-Ct samples substantially improved the sequencing results from these samples. Of the 324 samples tested in the high Ct range of 26.5 to 33.5, we observed 109 samples which reconstructed without hybrid capture compared to 181 samples that reconstructed with the application of hybrid capture (**Figure 4D**). We also observed a longer average genome length (**Figure 4E**) and fewer uncalled bases (**Figure 4F**) when hybrid capture was applied. Hybrid capture of high Ct samples achieved this by yielding both higher total reads and, more importantly, higher on-target reads for these samples. We were also able to improve the processing time of hybrid capture by testing the need of a dry-down procedure using SpeedVac as suggested by the Twist protocol. No compromise in quality of the final products was observed by replacing the concentrating step with another bead purification step **(Supplementary Figure 9)**.

## DISCUSSION

Here, we describe the development and optimization of a SARS-CoV-2 genome surveillance infrastructure for the city of New York. While all components of the pipeline underwent optimization, the greatest gains in genome reconstruction rate were observed from improvements in sample handling and transport, optimization of the RT-PCR reaction to convert viral RNA into cDNA, and strategies to improve reconstruction of high Ct samples, like inclusion of PEG in the tagmentatoin reaction and hybrid capture of prepared libraries. Reconstruction rates have improved to 85-95% for samples Ct < 30 and we can achieve appreciable reconstruction rates for samples up to Ct 34 as measured with the N1 probe (**Figure 5**). Both the diagnostic and sequencing pipelines are modular and scalable, enabling either local facilities to diagnose and sequence pathogens, or the aggregation of samples to a regional facility which serves a broader area for pathogen genome surveillance. Furthermore, this pipeline is adaptable to any DNA or RNA virus, requiring only development and optimization of the genome amplification step to plug into the current sequence methodology. For instance, we have developed the laboratory and analysis capabilities to sequence both influenza A and B for future surveillance efforts^28–30^,and anticipate developing the capability to sequence other pathogens.

**Figure 5.**
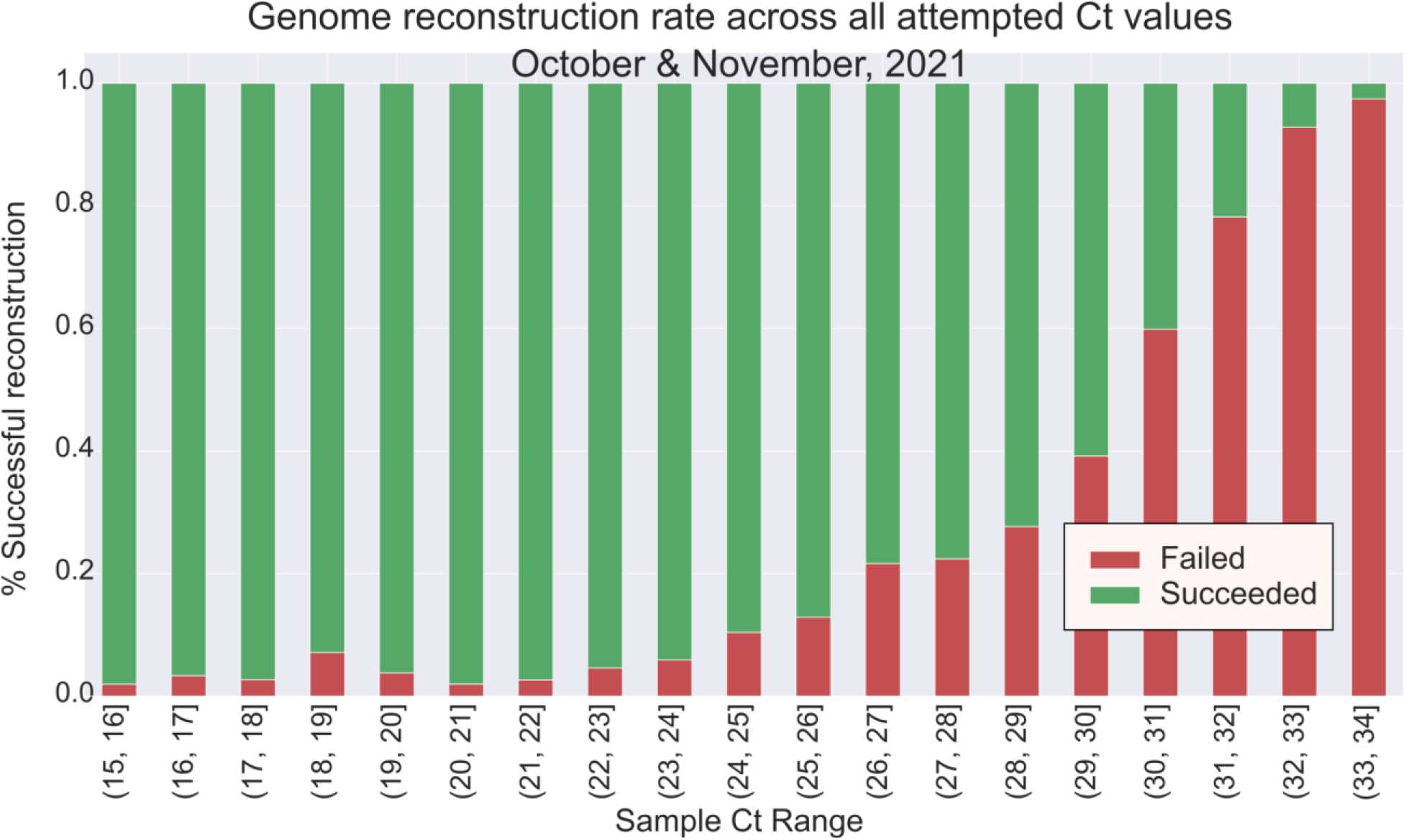
SARS-CoV-2 genome reconstruction rates across all attempted Ct values for the months of October and November, 2021. The cumulative effect of all the pipeline optimizations described here allow genome reconstruction for samples up to Ct = 34. In total, ∼87% of samples under Ct 30 were reconstructed in this 2-month period. Sample size = 7466.

The SARS-CoV-2 genome sequencing facility at Pandemic Response lab was a critical resource for preparing both the city of New York and the nation as a whole for the changing tides of the pandemic in the US. PRL has tracked the spread of the most consequential variants throughout NYC (**Figure 6**), enabling the public health apparatus of the city to predict the severity of new waves transmission and prepare the public health response. PRL was the first facility to detect the Omicron variant in New York State (4 cases reported coincident with New York City DOHMH Public Health Laboratory on December 2, 2021) and one of the earliest nationwide, and the rapid increase in frequency of this variant observed by our facility and others allowed the city to prepare for an unprecedented wave in transmission and the associated strains on the hospital system. As the events in NYC have served as an early warning for the rest of the US throughout the pandemic, the data gathered on variant spread here have been a key resource for decision making throughout the rest of the nation as well. Thus, a robust and continuing pathogen genomic surveillance system is mparticular, amplicons 5, 15, 21, aiportant in NYC, and in the other major cities where large scale sequencing operations have been built to surveil this pandemic, as an early warning system of the entry of possible new pandemic strains into the US^31–33^.

**Figure 6.**
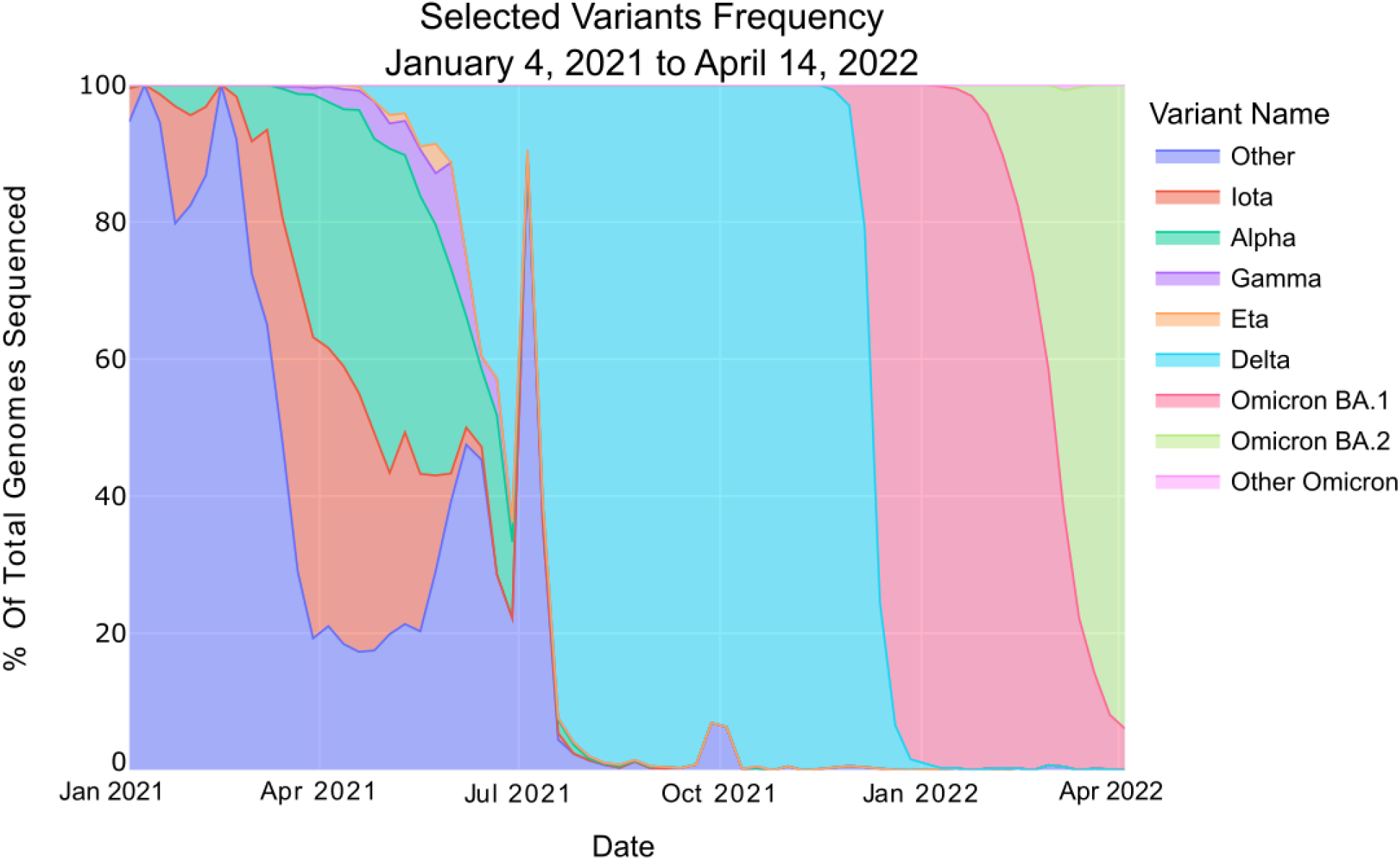
City-wide genomic surveillance enables tracking of strain prevalence over time. **(A)** Prevalence of various variants of concern (VOCs) throughout New York City from January 4, 2021 until April 14, 2022.

The ability to link our genomic surveillance data to patient demographics is also an important capability to measure the spread of new strains and the varying public health burden and socioeconomic impact of the pandemic on different demographic groups in the city. In particular, deidentified patient zip code information has enabled tracking of the origin and spread of new viral strains such as Omicron in near-real time (**Figure 7A-D**). This level of data granularity can help public health officials understand which communities are being impacted most and measure the effectiveness of neighborhood-level interventions. It has been well documented that the COVID-19 pandemic has hit low-income neighborhoods and communities of color in cities throughout the U.S. the hardest. The potential of this kind of high-resolution data marrying pathogen spread with geographic and demographic data is only just beginning to be realized, and has an important role to play in improving the equity of access to public health resources in our urban areas.

**Figure 7.**
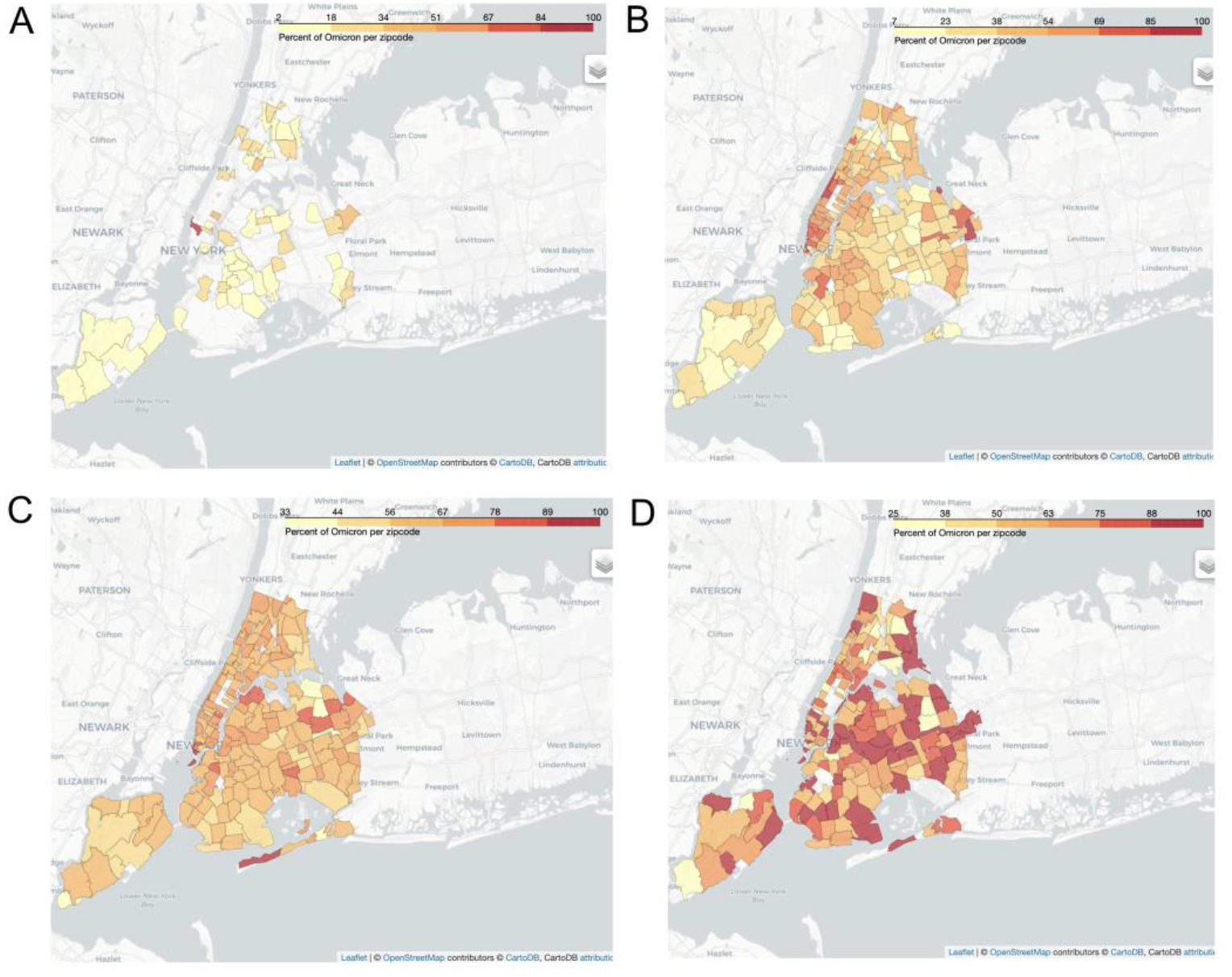
Frequency of the Omicron strain in SARS-CoV-2 cases during the week of December 2021. The prevalence of the Omicron strain is depicted by zip code in New York City for samples collected the weeks of **(A)** December 1-7th, **(B)** December 8-15th, **(C)** December 16-23rd, and **(D)** December 24-31st, 2021.

The COVID-19 crisis has highlighted the role of preparedness as the most critical element of pandemic response, and we anticipate that the tools built during this time will be essential to mitigate the likelihood and severity of future pandemics. We believe the utility of this pipeline in the fight against the COVID-19 pandemic provides a strong argument for broad adoption of genomic surveillance of this and other pathogens with the potential to spawn new pandemics which threaten public life and commerce. To safeguard against future public health threats, we anticipate that urban centers will elect to establish a localized and vertically integrated organization capable of city-scale qPCR testing and genome sequencing. As pandemic response gives way to pandemic prevention after the COVID-19 pandemic finally subsides, consistent genomic surveillance of circulating viral strains in our most dense urban centers will be among the most important tools to enable rapid proactive measures to prevent another crisis of this kind.

## MATERIALS AND METHODS

### RNA transport, storage, and hitpicking

Following testing at PRL-NYC, RNA plates containing SARS-CoV-2 positive samples with Ct of 34 and below are identified and barcoded for hitpicking. Using Tecan Fluent Automation Workstation, positive samples are hitpicked using custom hitpicking scripts to reformat all positive samples into a single 384-well PP Echo plate ordered by Ct, with the lowest Ct sample in A1 and ascending row by row. In addition, a Twist Synthetic Viral RNA positive control is added at wells B2 and P22, and a negative control in well P24, such that quadrant 4 of a given plate can be tested for quality control and contain both positive controls and the negative control, along with 95 test samples of increasing Ct. Hitpicked plates are kept at -80°C until transport to the R& D facility on dry ice. Upon arrival, plates are thawed on ice and used as template in a genomic RT-PCR on the same day. To confirm that these methods preserve RNA quality, RT-qPCR using both the N1 and N2 primer-probe sets are performed for each plate, and the values obtained are compared to those reported at the PRL testing facility.

### RNA Extraction of STAT samples at LIC R&D site

Reagent plates including binding plate, wash plate, 80% ethanol plate, and elution plate are prepared in advance for RNA extraction using ThermoScientific KingFisher Flex instrument. Binding plates are prepared by mixing 33 mL of pre-made Teknova Binding Solution (4M Guanidine Thiocyanate Buffer, 10 mM Tris-HCl, 1mM EDTA, 20% PEG 8000, 5% Tween-20, storage at 25 °C) with well-vortexed SpeedBead Magnetic Particles in a 50 mL Falcon tube. The tube is inverted five times to ensure even distribution of beads and then set on a rocker for 5 minutes. 300 uL of the final solution is dispensed into each well of a KingFisher 96 deep well plate and stored at 25 °C until use. The wash plate and ethanol plates are prepared by adding 500 uL of Teknova Wash Solution and 80% ethanol solution, respectively, into each well of KingFisher 96 deep well plates and then stored at 25 °C. Elution plates are prepared by dispensing 50 uL of TE Buffer (pH 8.4) into each well of a KingFisher 96 Well (200 uL) plate and stored at 25 °C. After reagent preparation, the biosafety cabinet work surface is thoroughly cleaned with Eliminase and 70% ethanol, and 200 uL of Proteinase K-treated patient samples are carefully transferred into each well of the binding plate and pipetted thoroughly with the binding solution. The sample plate and the reagent plates are placed accordingly on the KingFisher following the instructions of in-house protocol created for the RNA extraction, and a fresh set of tip comb was loaded before starting the protocol. After the run is completed, the extracted RNA sample plate is sealed immediately and stored in a 2 °C - 8 °C refrigerator until RT-PCR reaction on the same day.

### Sequencing STAT samples

A separate STAT pipeline was established at LIC in order to process high priority patient swabs sent directly from hospital emergency departments. The STAT samples are accessioned, reformatted into 96-well plate, and extracted using Kingfisher Flex (Thermo Scientific). Extracted RNA samples are then reformatted again into a 384-well plate using Cybio Felix (Analytik Jena), and the rest of the sequencing process is resumed following the protocol described for NYC Line samples.

### A1200 amplicon RT-PCR of hitpicked positive RNA plates

Mastermix plates are preassembled containing 5 μL of Takara One-Step Prime Script III 2x Mastermix in Thermo Scientific Armadillo clear 384-well PCR plates and stored at -20 °C. For each plate of positive hitpicked samples, 1 or 3 μL of SARS-CoV-2 RNA and H_2_O to 10 μL is pipetted into two Mastermix plates using the Analytick Jena CyBio Felix robot. These plates are then transferred to the Echo 525 acoustic pipettor, where 50 nL of 100 uM P1 or P2 A1200 Midnight primer mix^11^is dispensed to each well. Plates are then briefly spun down and cycled in Eppendorf Mastercycler X50t thermal cyclers using the following protocol optimized for our process: Reverse transcription reaction at 52 °C for 30 min followed by 35 cycles of 15 seconds denaturation at 95 °C, 3 minutes extension at 61 °C and a final cooling to 12 °C. Lots of Takara One-Step Prime Script III 2x Mastermix are routinely checked for efficacy, and enzyme is kept on had to revert to the 2-step RT-PCR method dictated by the Midnight protocol when necessary.

### Tagmentation of A1200 amplicons

Illumina Tagment Mix is prepared by creating a master mix containing 0.25 uL of Tagmentation Enzyme with 1.25 μL Tagment buffer for each well. The mix is dispensed into Thermo Scientific Armadillo clear 384-well PCR plates using Formulatrix Mantis and placed on ice. Following RT-PCR, 2 μL of A1200 P1 and 2 uL of P2 reactions are pooled together and diluted 30-fold into 120 uL of nuclease-free water. Samples are again Qubited and tagmented or diluted further to bring all samples under 0.5 ng/μL for efficient tagmentation. 1 μL of the diluted mixture is transferred to the tagmentation plate containing 1.5 uL Tagment Mix, and then placed in the thermocycler for tagmentation reaction. Tagmentation protocol is as follows: Thermocycler pre-heated to 55 °C before insertion of the plate, then tagmentation at 55 °C for 10 minutes.

### Barcoding of tagmented DNA and library amplification

Barcoding MasterMix is prepared by mixing 5 uL of 2X Kapa Hifi HotStart ReadyMix with 2.25 uL of nuclease-free water for each well. Following tagmentation, Kapa Mix is dispensed directly onto the tagmentation plate using MANTIS with high-volume chips. Using Echo 525 acoustic dispenser, 400 nL of 100 uM n7xx and 400 nL of 100 uM of n5xx oligo primers (Illumina) are dispensed consecutively onto the sample plate. Following addition of the primers, the barcoded samples - now 10 uL total volume - are placed on the thermocycler for barcoding PCR reaction. The optimized barcoding PCR reactions are as follows: 72 °C for 5 minutes, 98 °C for 5 minutes, 13 cycles of 10 sec of denaturation at 98 °C, 30 sec of annealing at 66 °C, and 30 seconds of extension at 72 °C. Final extension is done for 5 min at 72 °C before cooling to 10 °C. For each of the 384-well plates, the final libraries are categorized as either low Ct (Ct < 27) or high Ct (Ct > 27). 2 uL from each library is pooled into its respective low-bind tubes. All samples are purified using DNA Clean & Concentrator-5 (Zymo Research) and eluted in approximately 50 uL of elution buffer.

### Hybrid capture of high Ct samples

Hybrid capture is applied to high Ct (> 27) samples for target enrichment and better genome reconstruction. After pooling in equal volume and column purification, this pooled sample is further purified with a 1X volume ratio of AMPure beads and eluted in diH2O. Hybrid capture is then carried out using the Twist Target Enrichment Workflow and Twist SARS-CoV-2 Research Panel (Catalog# 102017) with minor adjustments to fit into our existing pipeline and the library construction method. The target-enriched library is then combined with the low-Ct samples from the same plate (see below) for sequencing.

### Library pooling and loading onto Illumina sequencer

Low Ct (< 27) samples from each plate are pooled in equal volumes and purified with DNA Clean & Concentrator-5 (Zymo Research) while high Ct (> 27) undergo hybrid capture. Following hybrid capture, the low and high Ct pools are combined in a ratio targeting ∼1 million reads per high Ct sample and ∼300,000 reads per low Ct sample to encourage reconstruction of poorer quality samples. DNA concentration of the pooled libraries are measured using Qubit 1X dsDNA HS Assay Kit and a BioTech Synergy H1 plate reader, low- and high-Ct pools are combined, and the sample is diluted to 4 nM. Libraries are then denatured and diluted according to the Denature and Dilute Libraries Guide by Illumina for MiSeq and NextSeq before being loaded on an appropriate Illumina sequencer.

### Genome assembly and variant calling

For each specimen, sequencing adapters are first trimmed using Trim Galore v0.6.6^34^, then aligned to the SARS-CoV-2 Wuhan-Hu-1 reference genome (NCBI Nucleotide NC_045512.2) using BWA MEM 0.7.17-r1188^35^. Reads that are unmapped or those that have secondary alignments are discarded. Consensus and mutations were called using samtools^36^and Intrahost variant analysis of replicates (iVar)^37^with a minimum quality score of 20, frequency threshold of 0.6 and a minimum read depth of 10x coverage. A consensus genome with ≥ 90% breadth-of-coverage with ≤ 3000 ambiguous bases is considered a successful reconstruction (as per APHL recommendation). For data reporting, variants are called using the most recent version of PANGOLIN^38^at the time of data reporting. PLEARN-v1.2.105 was used to call strains that were used to generate the plots in this manuscript.

### Generation of maps depicting geographical distribution of Omiron in NYC

Variant data obtained from our genomic sequencing pipeline was merged with geographic patient data from our clinical lab. The data was grouped by day, variant, and zip code. A Folium Choropleth map (https://python-visualization.github.io/folium/) was generated for each week of December where zipcodes are coloured based on the prevalence of Omicron samples in that zipcode for the given timeframe.

## Data Availability

All data produced in the present study are available upon reasonable request to the authors.

## ACKNOWLEDGMENTS

Thank you to Kenra Ford, Jen Rakeman-Cagno, Scott Hughes, Jade Wang, Andrea DeVito, Emma Claye, Jay Varma, Jennifer Gaumgartner, Elizabeth Luoma, and the many members of HHC, DOHMH, and De Blasio’s NYC Mayoral office for insightful discussions and close collaboration to build insightful data reporting. We would like to thank Vaishali Hodel for accommodating hitpicking on the PRL clinical line, and all individuals who helped with hitpicking positive samples, including Maxwell Shih, Merle Becker, David Ichikawa, Wolfgang Ott, Cierra Sing, Celeste Tompkins, Carmen Urgiles, Scarlett Sims, Erica Briggs, Minh Nguyen, Fallon Schwurack, Sylvia Zabielski, Fathma Abdalla, Rudy Rodriguez, Santi Jimenez, Ulysses Gonzalez, Rubi Singh, and Aliyah Subia. Thank you to Andrew Martin, Danielle Bulaon, AJ Cooper, and JT Lukens for help automating various processes. Thank you to Terra McNary, Alexa Holcombe, and Chen Wang for project and lab management of the sequencing division of PRL.

## AUTHOR CONTRIBUTIONS

HHL conceived the need for a PRL genome surveillance pipeline and proposed the initial experimental pipeline design. MJH, HHL, HH, and JL prototyped initial workflows. MJH and HH oversaw implementation, modification, and optimization of the wet lab sequencing pipeline. MJH, HH, and HHL trained the lab sequencing team on the wet lab pipeline. MJH, SCK, HH, CDC, MH, PB, SR, AC, DL, and KN optimized various components of the laboratory pipeline. HHL, IFE, WW, S. Chase, and S. Chhabria analyzed genome sequences and built informatic solutions for reporting. HHL oversaw the sequencing division of PRL prior to October 2021. MJH, SCK, HHL, JL, HH, IFE, and WW wrote the manuscript. MJH, WW, SCK, HH, and IFE generated figures. JL is the Director of R&D at PRL. All authors read and edited the manuscript.

## REFERENCES

1. Thompson CN, Baumgartner J, Pichardo C, et al. COVID-19 Outbreak — New York City, February 29–June 1, 2020. MMWR Morb. Mortal. Wkly. Rep. 69, 1725–1729 (2020).

2. Liu, Y. et al. Delta spike P681R mutation enhances SARS-CoV-2 fitness over Alpha variant. bioRxiv (2021) doi:10.1101/2021.08.12.456173.

3. Peacock, T. P. et al. The SARS-CoV-2 variants associated with infections in India, B.1.617, show enhanced spike cleavage by furin. bioRxiv 2021.05.28.446163 (2021) doi:10.1101/2021.05.28.446163.

4. Volz, E. et al. Evaluating the Effects of SARS-CoV-2 Spike Mutation D614G on Transmissibility and Pathogenicity. Cell 184, 64–75.e11 (2021).

5. Kistler, K., Huddleston, J. & Bedford, T. Rapid and parallel adaptive mutations in spike S1 drive clade success in SARS-CoV-2. bioRxiv (2022) doi:10.1101/2021.09.11.459844.

6. Lentini, A., Pereira, A., Winqvist, O. & Reinius, B. Monitoring of the SARS-CoV-2 Omicron BA.1/BA.2 variant transition in the Swedish population reveals higher viral quantity in BA.2 cases. bioRxiv (2022) doi:10.1101/2022.03.26.22272984.

7. Hirabara, S. M. et al. SARS-COV-2 Variants: Differences and Potential of Immune Evasion. Front. Cell. Infect. Microbiol. 11, 781429 (2021).

8. Liu, H., Wei, P., Kappler, J. W., Marrack, P. & Zhang, G. SARS-CoV-2 Variants of Concern and Variants of Interest Receptor Binding Domain Mutations and Virus Infectivity. Front. Immunol. 13, 825256 (2022).

9. Shu, Y. & McCauley, J. GISAID: Global initiative on sharing all influenza data - from vision to reality. Euro Surveill. 22, (2017).

10. Freed, N. E., Vlková, M., Faisal, M. B. & Silander, O. K. Rapid and inexpensive wholegenome sequencing of SARS-CoV-2 using 1200 bp tiled amplicons and Oxford Nanopore Rapid Barcoding. Biol Methods Protoc 5, (2020).

11. Freed, N. & Silander, O. SARS-CoV2 genome sequencing protocol (1200bp amplicon ‘midnight’ primer set, using Nanopore Rapid kit). https://www.protocols.io/view/sars-cov2-genome-sequencing-protocol-1200bp-amplic-bwyppfvn (2021) xdoi:10.17504/protocols.io.bwyppfvn.

12. Pillay, S. et al. Whole Genome Sequencing of SARS-CoV-2: Adapting Illumina Protocols for Quick and Accurate Outbreak Investigation during a Pandemic. Genes 11, (2020).

13. Jacot, D., Pillonel, T., Greub, G. & Bertelli, C. Assessment of SARS-CoV-2 Genome Sequencing: Quality Criteria and Low-Frequency Variants. J. Clin. Microbiol. 59, e0094421 (2021).

14. Coolen, J. P. M. et al. SARS-CoV-2 whole-genome sequencing using reverse complement PCR: For easy, fast and accurate outbreak and variant analysis. J. Clin. Virol. 144, 104993 (2021).

15. Sint, D., Raso, L. & Traugott, M. Advances in multiplex PCR: balancing primer efficiencies and improving detection success. Methods Ecol. Evol. 3, 898–905 (2012).

16. Nguyen-Dumont, T. et al. Cross-platform compatibility of Hi-Plex, a streamlined approach for targeted massively parallel sequencing. Anal. Biochem. 442, 127–129 (2013).

17. Pipelines R&D, D. N. A. et al. COVID-19 ARTIC v3 Illumina library construction and sequencing protocol v5. protocols.io (2020) doi:10.17504/protocols.io.bibtkann.

18. Ma, W. et al. Genomic perspectives on the emerging SARS-CoV-2 Omicron variant. Genomics Proteomics Bioinformatics (2022) doi:10.1016/j.gpb.2022.01.001.

19. Picelli, S. et al. Tn5 transposase and tagmentation procedures for massively scaled sequencing projects. Genome Res. 24, 2033–2040 (2014).

20. Adey, A. et al. Rapid, low-input, low-bias construction of shotgun fragment libraries by high-density in vitro transposition. Genome Biol. 11, R119 (2010).

21. Gertz, J. et al. Transposase mediated construction of RNA-seq libraries. Genome Res. 22, 134–141 (2012).

22. Lu, B. et al. Transposase-assisted tagmentation of RNA/DNA hybrid duplexes. Elife 9, (2020).

23. Di, L. et al. RNA sequencing by direct tagmentation of RNA/DNA hybrids. Proc. Natl. Acad. Sci. U. S. A. 117, 2886–2893 (2020).

24. Mamanova, L. et al. Target-enrichment strategies for next-generation sequencing. Nat. Methods 7, 111–118 (2010).

25. Machyna, M. & Simon, M. D. Catching RNAs on chromatin using hybridization capture methods. Brief. Funct. Genomics 17, 96–103 (2018).

26. Nagy-Szakal, D. et al. Targeted Hybridization Capture of SARS-CoV-2 and Metagenomics Enables Genetic Variant Discovery and Nasal Microbiome Insights. Microbiol Spectr 9, e0019721 (2021).

27. Gaudin, M. & Desnues, C. Hybrid Capture-Based Next Generation Sequencing and Its Application to Human Infectious Diseases. Front. Microbiol. 9, 2924 (2018).

28. Zhou, B. et al. Universal influenza B virus genomic amplification facilitates sequencing, diagnostics, and reverse genetics. J. Clin. Microbiol. 52, 1330–1337 (2014).

29. Hoffmann, E., Stech, J., Guan, Y. & Webster, R. G. Universal primer set for the full-length amplification of all influenzaA viruses. Arch. Virol. 146, 2275–2289 (2001).

30. Borges, V., Pinheiro, M., Pechirra, P., Guiomar, R. & Gomes, J. P. INSaFLU: an automated open web-based bioinformatics suite ‘from-reads’ for influenza whole-genome-sequencing-based surveillance. Genome Med. 10, 46 (2018).

31. Long, S. W. et al. Molecular Architecture of Early Dissemination and Massive Second Wave of the SARS-CoV-2 Virus in a Major Metropolitan Area. MBio 11, (2020).

32. Lemieux, J. E. et al. Phylogenetic analysis of SARS-CoV-2 in Boston highlights the impact of superspreading events. Science 371, (2021).

33. Robishaw, J. D. et al. Genomic surveillance to combat COVID-19: challenges and opportunities. Lancet Microbe 2, e481–e484 (2021).

34. Krueger, F. Trim Galore!: A wrapper tool around Cutadapt and FastQC to consistently apply quality and adapter trimming to FastQ files. https://github.com/FelixKrueger/TrimGalore.

35. Li, H. & Durbin, R. Fast and accurate short read alignment with Burrows-Wheeler transform. Bioinformatics 25, 1754–1760 (2009).

36. Li, H. et al. The Sequence Alignment/Map format and SAMtools. Bioinformatics 25, 2078–2079 (06 2009).

37. Grubaugh, N. D. et al. An amplicon-based sequencing framework for accurately measuring intrahost virus diversity using PrimalSeq and iVar. Genome Biol. 20, 8 (2019).

38. O’Toole, Á. et al. Assignment of epidemiological lineages in an emerging pandemic using the pangolin tool. Virus Evol 7, veab064 (2021).

