## Supplementary Material for "Development and scaling of a sequencing pipeline for genomic surveillance of SARS-CoV-2 in New York City"

[1] Research and Development, Pandemic Response Lab.
30-02 48th Ave. Long Island City, NY 11101

‡ Equal Contribution
* To whom correspondence should be addressed. Email:
,

[†] Present address: Cultivarium, Boston, MA.
[♀] Present address: Kern Systems, Allston, MA.

Keywords:
SARS-CoV-2, genomic surveillance, multiplexed RT-PCR, COVID-19, Variants of Concern, Viral Sequencing

### SUPPLEMENTARY MATERIAL

### SUPPLEMENTARY TABLES

| Sample number | N1 Ct | genome_length | n_count | reconstruction | startpos | endpos | numreads | % reads | covbases | coverage | average_coverage | meanbaseq | meanmapq | per_GeneLength |
|---|---|---|---|---|---|---|---|---|---|---|---|---|---|---|
| 1 | 36.61 | 28692 | 27240 | FALSE | 1 | 29870 | 636174 | 0.849633499 | 28204 | 94.4225 | 507.253 | 34 | 42.5 | 96.05624372 |
| 2 | 16.45 | 29763 | 0 | TRUE | 1 | 29870 | 70460153 | 94.10209521 | 29863 | 99.9766 | 174950 | 34.8 | 60 | 99.64178105 |
| 3 | 36.78 | 27908 | 27110 | FALSE | 1 | 29870 | 94769 | 0.126567444 | 17562 | 58.7948 | 75.8203 | 33.9 | 39.7 | 93.43153666 |
| 4 | 34.05 | 29745 | 28305 | FALSE | 1 | 29870 | 638308 | 0.852483533 | 10988 | 36.7861 | 582.205 | 34.2 | 42.2 | 99.58151992 |
| 5 | 29.58 | 29754 | 26013 | FALSE | 1 | 29870 | 1303975 | 1.741506006 | 15170 | 50.7867 | 2326.9 | 34.6 | 53.5 | 99.61165049 |
| 6 | 40 | 29759 | 28458 | FALSE | 1 | 29870 | 284714 | 0.380245895 | 16630 | 55.6746 | 224.981 | 33.9 | 41.2 | 99.62838969 |
| 7 | 37 | 29741 | 28382 | FALSE | 1 | 29870 | 379046 | 0.506229709 | 13782 | 46.1399 | 327.514 | 33.9 | 40.7 | 99.56812856 |
| 8 | 35.97 | 28691 | 24311 | FALSE | 1 | 29870 | 1078926 | 1.440944887 | 15596 | 52.2129 | 1801.53 | 34.6 | 51.5 | 96.05289588 |

**Supplementary Table 1: Over-sequencing of high-Ct SARS-CoV-2 positive samples fails to rescue reconstruction.** Seven samples with high Ct and one with low Ct (Sample 2, Ct = 16.45) were sequenced on an Illumina NextSeq 500/550 Mid-Output v2.5 Kit (150 cycles), representing a substantial excess of sequencing reads necessary to reconstruct these genomes. Despite this over-sequencing of the samples, all high-Ct samples failed to reconstruct. They also suffered a high number of undetermined bases (n_count).

| | 1 uL | 5 uL |
|---|---|---|
| TRUE | 18 | 25 |
| FALSE | 19 | 12 |
| Saved | 7 | |

($n$ = 37)

**Supplementary Table 2: Increasing template input volume improves genome reconstruction of SARS-CoV-2 positive samples**. To assess the impact of input volume on reconstruction rate in our pipeline, 37 independent SARS-CoV-2 positive patient samples were sequenced each using 1 μL or 5 μL as template. Of these samples, 18 successfully reconstructed when using 1 μL and 25 reconstructed when using 5 μL.

### SUPPLEMENTARY FIGURES

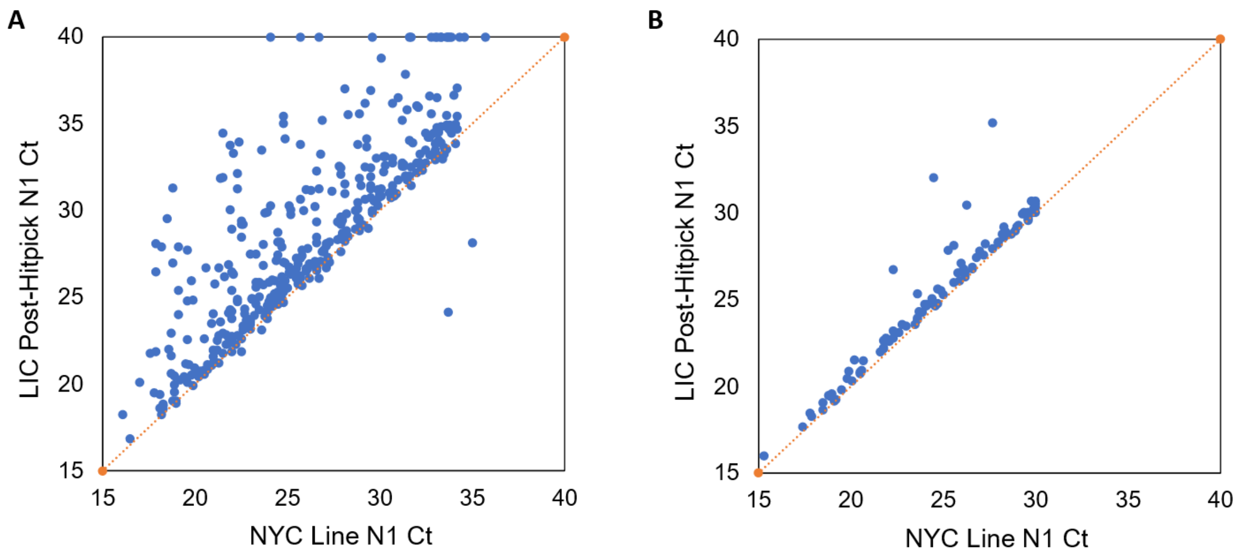


**Supplementary Figure 1: Effect of QC measures on RNA quality during transport.** Ct values are routinely measured for samples arriving at the sequencing facility after being diagnosed and hitpicked at the clinical lab. (A) Samples which have been degraded due to improper handling such as extended time at ambient temperature or multiple freeze-thaws can be identified by deviation from the Ct value obtained at the clinical lab, seen as scatterplot points which rise above the identity line. (B) Samples which are handled properly should deviate little if at all from the original Ct values.

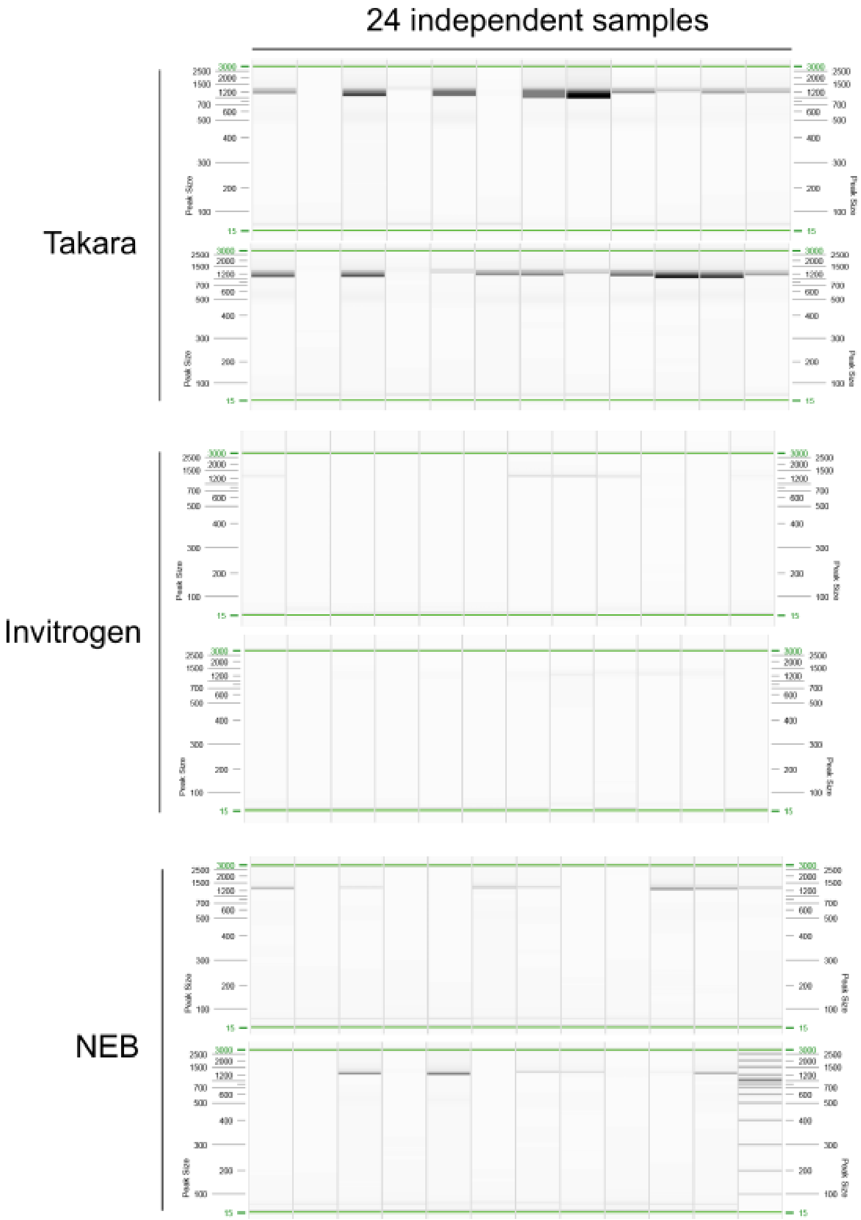


**Supplementary Figure 2.** Genomic PCR test of SARS-CoV-2 positive samples using 1-step RT-PCR kits. Takara One Step PrimeScript III (#RR600A), Invitrogen SuperScript™ IV One-Step (#12594025), and NEB OneTaq One-Step (#E5315S) kits were tested for their ability to produce genomic cDNA in an A1200 RT-PCR of 24 SARS-CoV-2 positive samples. Resulting cDNA was run on a Qiaxcel high sensitivity capillary gel electrophoresis cartridge. Takara outperformed all other kits, producing more robust bands in more samples than other kits.

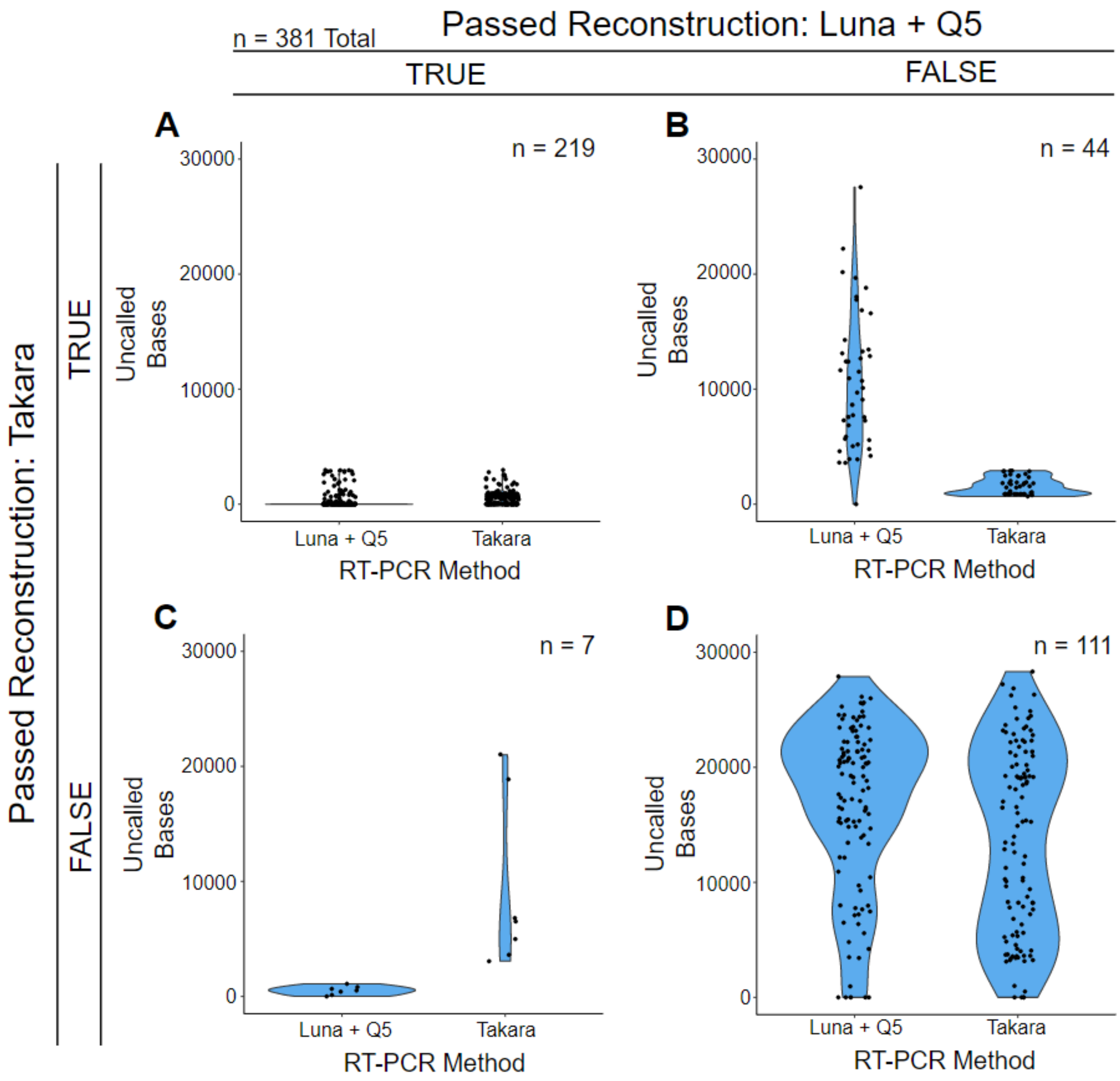


**Supplementary Figure 3: Violin plots of number of uncalled bases per genome for samples sequenced with Lunascript + Q5 versus Takara.** The two-step RT-PCR protocol is compared to the Takara one-step protocol. Plots are depicted in a truth table of whether samples were reconstructed in the two-step or one-step protocol. 44 samples reconstructed only using Takara, while only 7 samples reconstructed only using the two-step protocol. Notably, even in samples where neither kit was able to reconstruct the genome, the distribution of uncalled bases is shifted lower for the Takara kit relative to the two-step protocol.

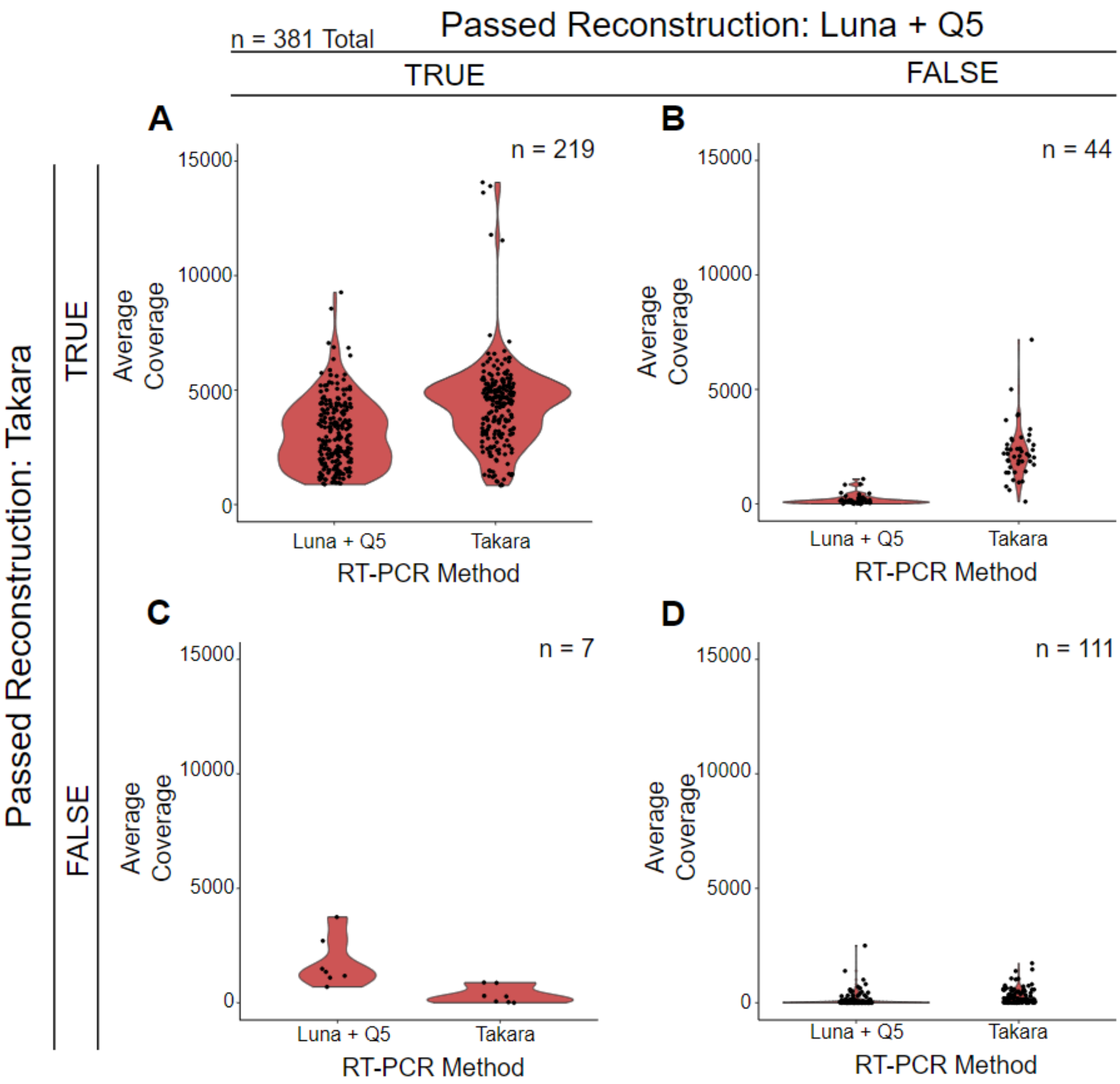


**Supplementary Figure 4: Average per sample coverage for samples sequenced with Lunascript + Q5 versus Takara.** The two-step RT-PCR protocol is compared to the Takara one-step protocol. Plots are depicted in a truth table of whether samples were reconstructed in the two-step or one-step protocol. 44 samples reconstructed only using Takara, while only 7 samples reconstructed only using the two-step protocol. Notably, for samples which were reconstructed using both kits, the average coverage is shifted greater in those reconstructed with the Takara kit, suggesting a greater proportion of on-target reads.

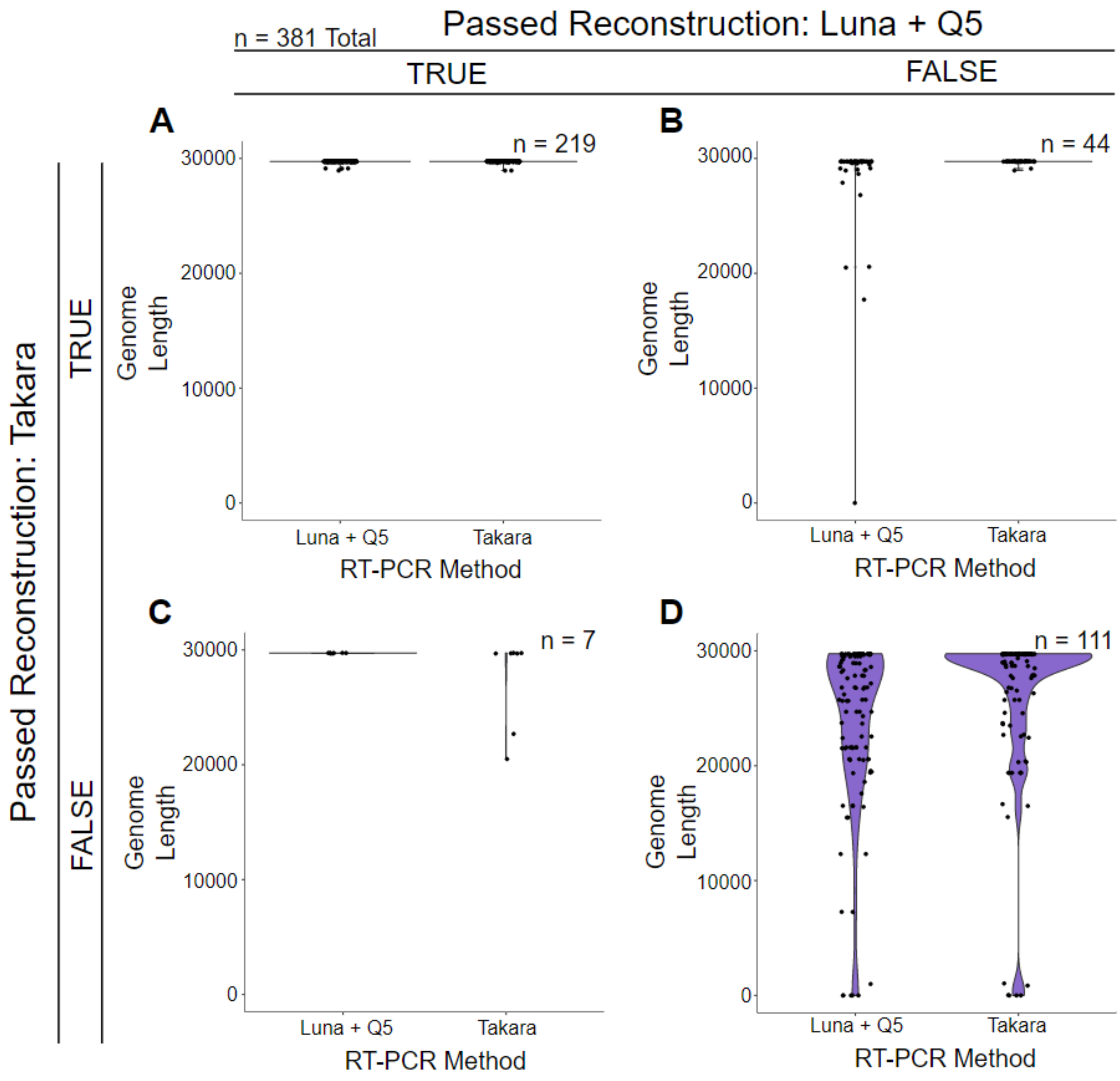


**Supplementary Figure 5: Genome length for samples sequenced with Lunascript + Q5 versus Takara.** The two-step RT-PCR protocol is compared to the Takara one-step protocol. Plots are depicted in a truth table of whether samples were reconstructed in the two-step or one-step protocol. 44 samples reconstructed only using Takara, while only 7 samples reconstructed only using the two-step protocol. Notably, even in samples where neither kit was able to reconstruct the genome, genome length is shifted longer for samples reconstructed using the Takara kit over the two-step protocol.

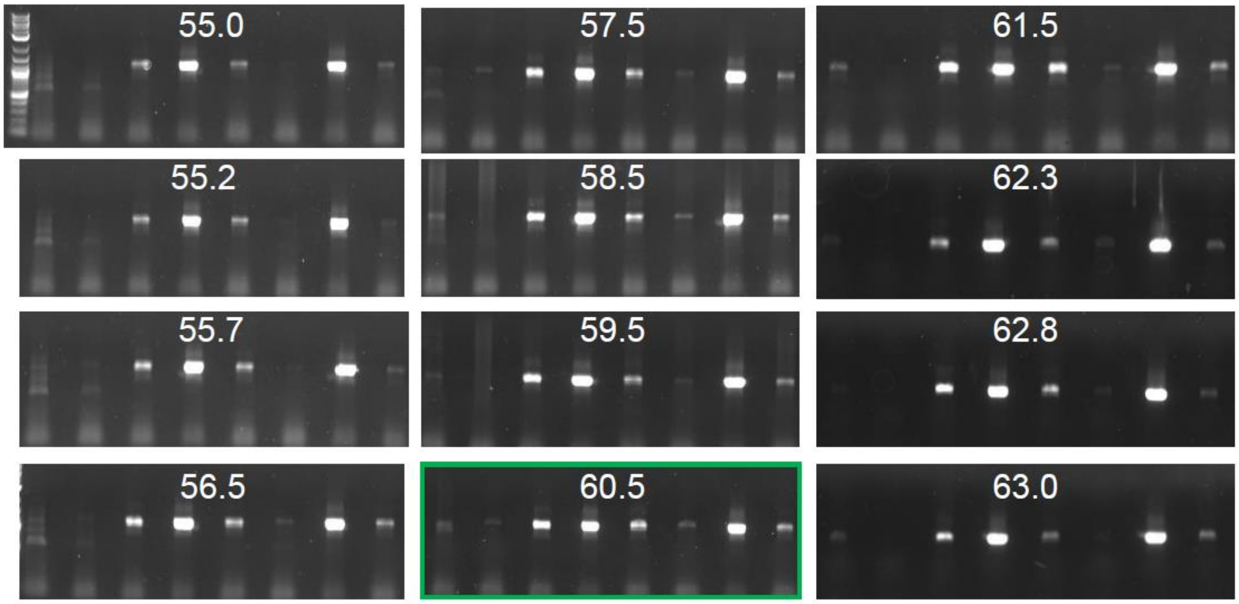


**Supplementary Figure 6: Elongation temperature optimization for RT-PCR of the SARS-CoV-2 genome.** The elongation temperature of the two-step Takara RT-PCR protocol was optimized using a gradient PCR on 8 independent SARS-CoV-2 positive samples. While the most robust samples succeed in all cases, faint bands for less ideal samples are more noticeable in lanes 1, 2, and 6 with fewer off products in the 60.5 degree elongation temperature condition.

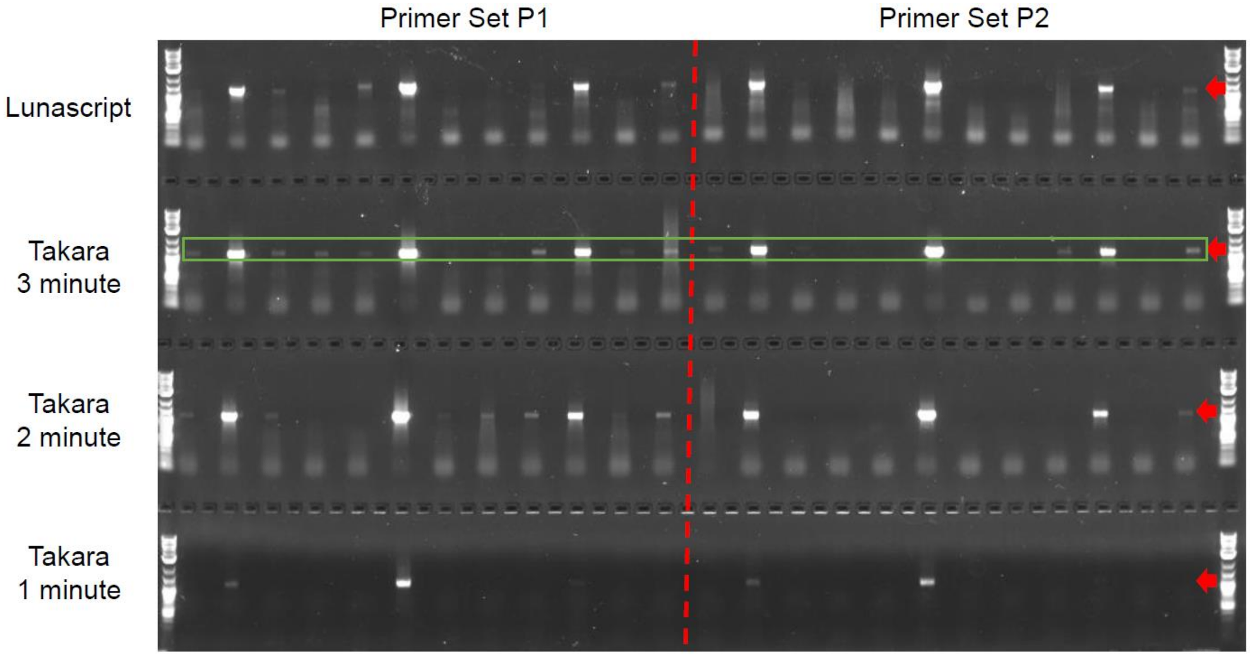


**Supplementary Figure 7: Elongation time optimization for RT-PCR of the SARS-CoV-2 genome.** The duration of the elongation step was optimized by performing RT-PCR with 1, 2, or 3 minute elongation steps on 12 independent SARS-CoV-2 positive samples. The 3 minute elongation time was found to produce robust cDNA amplicons in the most conditions without producing aberrant or large molecular weight off-products.

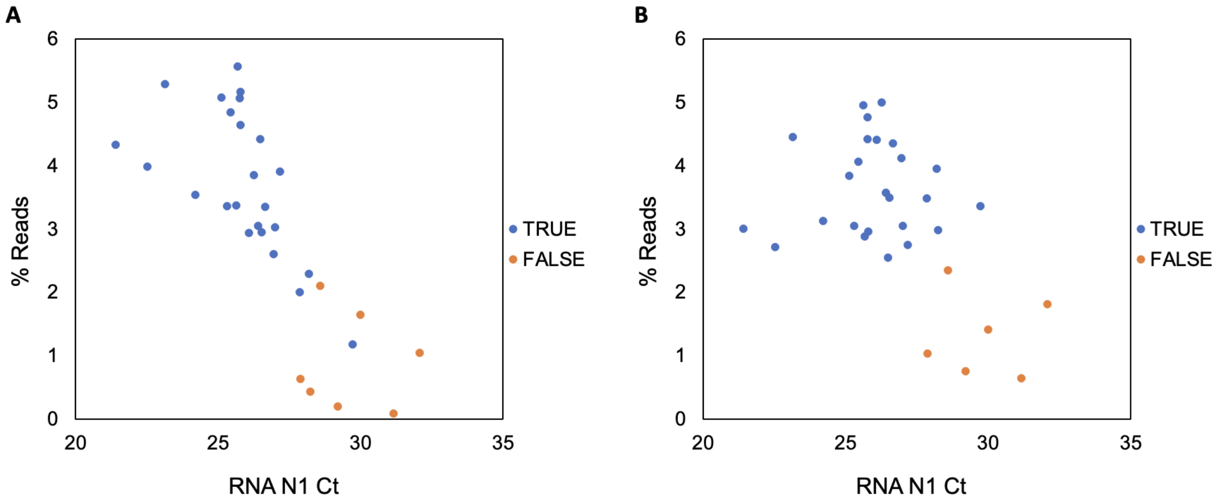


**Supplementary Figure 8. Effect of manual normalization on DNA libraries.**
Sequencing read depth versus RNA N1 Ct (20 < Ct < 32), *n*= 32 for **A** Unnormalized libraries and **B** Manually normalized libraries. (Blue = genome reconstructed, orange = genome failed to reconstruct)

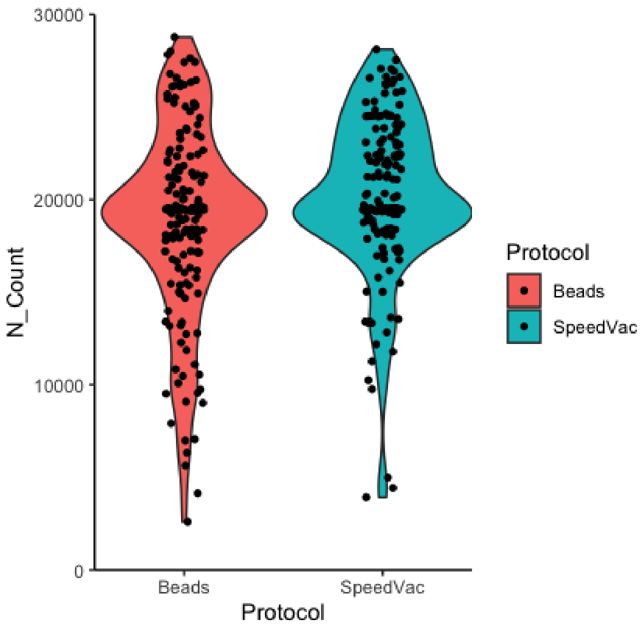


**Supplementary Figure 9: Elimination of SpeedVac step does not harm reconstruction of genomes undergoing hybrid capture.** To assess the need for a SpeedVac step in the hybrid capture protocol, 330 high-Ct (>30) samples were tested with and without the SpeedVac step. No significant difference was observed between samples undergoing these alternative treatments.
